# The Trend of Neutralizing Antibody Response Against SARS-CoV-2 and the Cytokine/Chemokine Release in Patients with Differing Severities of COVID-19: All Individuals Infected with SARS-CoV-2 Obtained Neutralizing Antibody

**DOI:** 10.1101/2020.08.05.20168682

**Authors:** Lidya Handayani Tjan, Tatsuya Nagano, Koichi Furukawa, Mitsuhiro Nishimura, Jun Arii, Sayo Fujinaka, Sachiyo Iwata, Yoshihiro Nishimura, Yasuko Mori

## Abstract

**Background:** COVID-19 patients show a wide clinical spectrum ranging from mild respiratory symptoms to severe and fatal disease, and older individuals are known to be affected more severely. Neutralizing antibody for viruses is critical for their elimination, and increased cytokine/chemokine levels are thought to be related to COVID-19 severity. However, the trend of the neutralizing antibody production and cytokine/chemokine levels during the clinical course of COVID-19 patients with differing levels of severity has not been established.

**Methods:** We serially collected 45 blood samples from 12 patients with different levels of COVID-19 severity, and investigated the trend of neutralizing antibody production using authentic SARS-CoV-2 and cytokine/chemokine release in the patients’ clinical courses.

**Results:** All 12 individuals infected with SARS-CoV-2 had the neutralizing antibody against it, and the antibodies were induced at approx. 4-10 days after the patients’ onsets. The antibodies in the critical and severe cases showed high neutralizing activity in all clinical courses. Most cytokine/chemokine levels were clearly high in the critical patients compared to those with milder symptoms.

**Conclusion:** Neutralizing antibodies against SARS-CoV-2 were induced at a high level in the severe COVID-19 patients, indicating that abundant virus replication occurred. Cytokines/chemokines were expressed more in the critical patients, indicating that high productions of cytokines/chemokines have roles in the disease severity. These results may indicate that plasma or neutralizing antibody therapy could be a first-line treatment for older patients to eliminate the virus, and corticosteroid therapy could be effective to suppress the cytokine storm after the viral genome’s disappearance.

## INTRODUCTION

In December 2019, a new coronavirus was identified as the cause of a pneumonia outbreak in Wuhan, China [1]. The identified virus was related to severe acute respiratory syndrome coronavirus (SARS-CoV), and it was thus named SARS-CoV-2 [2]. Since then, SARS-CoV-2 has spread to almost of the world’s countries. According to World Health Organization (WHO) data per 31 July 2020, there were more than 17 million confirmed cases with approx. 650,000 deaths worldwide.

The clinical spectrum of COVID-19 varies from mild to severe and fatal infections. The comorbidities related to disease severity include older age, hypertension, diabetes mellitus, obesity, malignancy, and more [3–5]. In addition to the identified comorbidities associated with poorer prognosis in COVID-19, unique immune responses against SARS-CoV-2 infection among different individuals may explain different clinical outcomes observed in infected populations. Patients with a severe infection induce a higher antibody titer compared to those with a mild infection[6], and this is also correlated with neutralizing antibody titer [6, 7]

Fatality due to COVID-19 has been reported to be linked to a hyperinflammmatory state characterized by the cytokine release syndrome (CRS) [8]. Among many different types of cytokines and chemokines that have been identified, interleukin (IL)-6 has been consistently reported to reflect the clinical severity of COVID-19 and has been the target of therapy in severe cases [9, 10].

Based on the available studies regarding the immune response against SARS-CoV-2, it is clear that humoral immunity is induced in COVID-19 patients. However, the trend of neutralizing antibody production and cytokine/chemokine levels during the clinical course of COVID-19 in patients with differing levels of disease severity has not been well explored. Here, we serially collected blood samples from COVID-19 patients with different disease severities and analyzed the trend of neutralizing antibody production during patients’ clinical courses by using authentic SARS-CoV-2 and determining the cytokine/chemokine release.

Although recent studies have used a pseudotyped virus for neutralizing assay[11, 12], inconsistent results in neutralizing assay have been shown between pseudotyped and authentic viruses [13–15]. In this study, authentic virus was consistently used to examine the neutralizing activity against SARS-CoV-2.

## PATIENTS AND METHODS

### Population and samples

A total of 12 COVID-19 patients, diagnosed both clinically and based on detection of virus genome by polymerase chain reaction (PCR) were included in this study. All patients were hospitalized at Hyogo Prefectural Kakogawa Medical Center, which is one of the 55 publicly designated medical institutions for infectious diseases in Japan. Blood samples were serially collected from each of the patients.

Additionally, seven age-matched healthy adults who were SARS-CoV-2 IgG-negative medical staff from the same facility we described [16] and Kita-harima Medical Center were included in this study as negative controls. IgG was examined using an immunochromatographic test kit (2019-nCoV Ab Test; INNOVITA, Hebei, China) as described [16]

### Serum neutralizing assay against SARS-CoV-2

To evaluate neutralizing activity of serum, we performed a neutralization test against SARS-CoV-2 (Biken-2 strain) which was kindly provided from Research Foundation for Microbial Diseases of Osaka University (BIKEN), Osaka University. The neutralization test was performed in biosafety level 3 laboratory. For this purpose, 4 x 10^4^ Vero E6 (TMPRSS2) cells [17] per well were seeded in 96-well tissue culture microplates 24 hr before the assay. Duplicate samples of a three-fold serial dilution of heat-inactivated (56°C, 30 min) serum were prepared using Dulbecco’s Modified Eagle’s Medium as the diluent and mixed with 100 tissue culture infectious dose (TCID)_50_ of virus and incubated at 37°C for 1 hr. After this incubation, the serum-virus mixture was added to Vero E6 (TMPRSS2) cells and incubated at 37°C for 3 days. Neutralizing antibody titer was determined as the highest serum dilution that did not show any cytopathic effects by observation under microscope. These experiments were repeated three times independently.

### Chemokine measurements

A total of 48 cytokines, chemokines, and growth factors were measured in serum samples using the Bio-Plex Pro Human Cytokine Screening 48-plex panel (Bio-Rad, Hercules, CA), and results were read using the Bio-Plex 200 system following the manufacturer’s instructions. The cytokines, chemokines, and growth factors detected in this system are listed in Table 1.

**Table 1.** List of cytokines, chemokines and growth factor analyzed in this study

|  |  |  |  |
| --- | --- | --- | --- |
| IL-1β | Eotaxin | IL-1α | MIF |
| IL-1ra | FGF basic | IL-2Rα | MIG |
| IL-2 | G-CSF | IL-3 | β-NGF |
| IL-4 | GM-CSF | IL-12p40 | SCF |
| IL-5 | IFN-γ | IL-16 | SCGF-β |
| IL-7 | IP-10 | CTACK | SDF-1α |
| IL-9 | MCP-1(MCAF) | GROα | TNF-β |
| IL-10 | MIP-1α | HGF | TRAIL |
| IL-12(p70) | PDGF-bb | IFN-α2 | IL-6 |
| IL-13 | MIP-1β | LIF | IL-8 |
| IL-15 | RANTES | MCP-3 | IL-18 |
| IL-17 | VEGF | M-CSF | TNF-α |

**IL**, interleukin; **FGF**, fibroblast growth factor; **G-CSF**, granulocyte colony-stimulating factor; **GM-CSF**, granulocyte-macrophage colony-stimulating factor; **IFN**, interferon; **IP**, interferon gamma-induced protein; **MCP**, monocyte chemotactic and activating factor; **MIP**, macrophage inflammatory protein; **PDGF**, platelet-derived growth factor; **RANTES**, regulated on activation, normal T-cell expressed and secreted; **VEGF**, vascular endothelial growth factor; **CTAK**, cutaneous T-cell-attracting chemokine; **GRO**, growth-related oncogene; **HGF**, hepatocyte growth factor; **LIF**, Leukemia inhibitory factor; **MCP**, monocyte chemotactic and activating factor; **M-CSF**, macrophage colony stimulating factor; **MIF**, macrophage migration inhibitory factor; **MIG**, monokine induced by interferon-gamma, **NGF**, nerve growth factor; **SCF**, stem cell factor; **SCGF**, stem cell growth factor; **SDF**, stromal cell-derived factor; **TNF**, tumor necrosis factor; **TRAIL**, TNF-related apoptosis-inducing ligand

### Ethics approval

The institutional review boards approved the study, and written informed consent was obtained from all participants.

## RESULTS

### Patient characteristics

Following ‘Interim Guidance, Clinical Management of COVID-19’ published by the WHO on 27 May 2020, one of the 12 patients was categorized as asymptomatic, one patient as mild, three patients as severe; and the remaining seven patients as critical infection. Eleven patients were >50 years old, and the remaining patient (K-Px-11, with a mild infection) was 40 years old. Five of the seven patients in critical-infection group were male, and three of these five patients have died as of this writing. The demographic and clinical data of patients are summarized in Table 2.

**Table 2.** Demographic and Clinical Characteristic of Patients

| Code | Sex, Age | Severity<sup>a</sup> | Medical history<sup>b</sup> | Hospitalization period (dpo)<sup>c</sup> | Upper Respiratory symptom<sup>d</sup> | Pneumonia | Persistent imaging abnormalities | Ventilator introduction (dpo)<sup>e</sup> | Ventilator removal (dpo)<sup>e</sup> | Treatment<sup>e</sup> | Equipment<sup>f</sup> | Outcome |
| --- | --- | --- | --- | --- | --- | --- | --- | --- | --- | --- | --- | --- |
| K-Px-10 | M, 59 | Asymptomatic | DM, glaucoma | 20 | - | N.D. | N.D. | - | - | - | - | Alive |
| K-Px-11 | M, 40 | Mild | - | 40 | + | - | - | - | - | - | - | Alive |
| K-Px-12 | M, 71 | Severe | AR, BA, BPH | 35 | + | + | + | - | - | C | - | Alive |
| K-Px-1 | F, 59 | Severe | Cervical cancer, HL, HT, SAS | 23 | + | + | + | - | - | C, F | - | Alive |
| K-Px-9 | F, 69 | Severe | HL, HT | 21 | + | + | + | - | - | C, F | - | Alive |
| K-Px-2 | M, 57 | Critical | - | 56 | + | + | + | 8 | 33 | F, L/R | CHDF, HD | Alive |
| K-Px-3 | M, 74 | Critical | DM, glaucoma, HL | 44 | + | + | + | 11 | 50 | C, F, L/R | CHDF | Dead (51 dpo<sup>e</sup>) |
| K-Px-4 | F, 63 | Critical | Osteoporosis | 80 | + | + | + | 16 | 65 | C, F, L/R | ECMO | Alive |
| K-Px-5 | M, 65 | Critical | HT, hyperuricemia | 93 | + | + | - | 12 | 32 | C, F, L/R | CHDF | Alive |
| K-Px-6 | M, 79 | Critical | Arrhythmia, CI, dementia | 16 | + | + | + | 6 | 15 | C, F, L/R | - | Dead (16 dpo<sup>e</sup>) |
| K-Px-7 | M, 78 | Critical | AAA, AP, arrhythmia, ASO, CHF, CKD, DM, GB, HT | 17 | + | + | + | 8 | 21 | C, F | - | Dead (22 dpo<sup>e</sup>) |
| K-Px-8 | F, 79 | Critical | DM, IgA nephropathy | 73 | + | + | + | 0 | 36 | F, L/R, Steroid (6-9 dpo<sup>e</sup>) | CRRT | Alive |
| H1 | F, 59 | Healthy | - | - | N.D. | N.D. | N.D. | - | - | - | - | - |
| H2 | M, 55 | Healthy | - | - | N.D. | N.D. | N.D. | - | - | - | - | - |
| H3 | M, 58 | Healthy | - | - | N.D. | N.D. | N.D. | - | - | - | - | - |
| H4 | M, 62 | Healthy | - | - | N.D. | N.D. | N.D. | - | - | - | - | - |
| H5 | F, 45 | Healthy | - | - | N.D. | N.D. | N.D. | - | - | - | - | - |
| H6 | F, 40 | Healthy | - | - | N.D. | N.D. | N.D. | - | - | - | - | - |
| H7 | M, 65 | Healthy | - | - | N.D. | N.D. | N.D. | - | - | - | - | - |

<sup>a</sup>. Severity criteria was as follows; **Mild**: upper respiratory symptoms without pneumonia ; **Moderate**: pneumonia requiring no supplemental oxygen; **Severe**: requirement for supplemental oxygen; **Critical**; requirement for intensive care including mechanical ventilation <sup>b</sup>. **AR**, allergic rhinitis; **BA**, bronchial asthma; **BPH**, benign prostate hyperplasia; **HL**, hyperlipidemia; **HT**, hypertension; **SAS**, sleep apnea syndrome; **DM**, diabetes mellitus; **CI**, cerebral infarction; **AAA**, abdominal aortic aneurysm; **AP**, angina pectoris; **ASO**, arteriosclerosis obliterans; **CHF**, chronic heart failure; **CKD**, chronic kidney disease; **GB**, gallbladder stone <sup>c</sup>. dpo stands for days post-onset of symptoms. <sup>d</sup>. Upper respiratory symptom included fever, cough and sore throat. <sup>e</sup>. **C**: ciclesonide, inhaled corticosteroid; **F**: favipiravir, broad-spectrum antiviral for influenza; **L/R**: lopinavir/ritonavir, oral antiviral for human-immunodeficiency virus; **Steroid**: Steroid pulse with 0.5 g methylprednisolone per day during the indicated period. <sup>f</sup>. **CHDF**, continuous hemodiafiltration; **HD**, hemodialysis; **ECMO**, extracorporeal membrane oxygenation; **CRRT**, continuous renal replacement therapy N.D.: Not determined

We define ‘COVID-19 critical patients’ as those who suffer from acute respiratory distress syndrome (ARDS) and require mechanical ventilation support. The duration of mechanical ventilation support for 7 patients with critical infection in this study ranged from 10 to 50 days, with shortest duration at 10 days in patient K-Px-6 who died after 16 days of hospitalization. In general, the durations of hospitalization in patients with a critical infection were longer compared to those of the other severity groups. There were two critically infected patients with relatively short durations of hospitalization (16 days for K-Px-6 and 17 days for K-Px-7), but these durations were short because the patients died at day 16 and day 17 after admission.

Among seven patients with critical disease, five had identified comorbidities related to COVID-19 clinical severity; another had a medical history unrelated to COVID-19 severity (osteoporosis), and the remaining patient did not have any background disease. All three patients who died were males >70 years old who had significant comorbidities related to COVID-19 disease severity. Of the three patients with severe disease, two had hypertension as a comorbidity related to COVID-19 severity. The mildly infected patient did not have any background disease. Of note, one asymptomatic patient had diabetes mellitus as a comorbidity.

### The neutralizing antibody titer’s trend in COVID-19 patients with different severities

Neutralizing antibody against virus is important for virus elimination. Therefore, to examine neutralizing antibody production for SARS-CoV-2, we serially collected blood samples from COVID-19 patients and determined the trend of neutralizing antibody titer against SARS-CoV-2 in serum of each patient.

A total of 45 blood samples from 12 patients were subjected to the measurement of neutralizing antibody activity. As shown in Figure 1, neutralizing antibody titer against SARS-CoV-2 was detected in all 12 COVID-19 patients regardless of their disease severity, whereas neutralizing antibody was not detected at all in healthy controls. The neutralizing antibodies in 10 patients with severe or critical disease showed relatively high titers compared to the asymptomatic patient (K-Px-10), although patient with mild symptoms (K-Px-11) also showed a high titer measured at day 42 after onset; this patient was a 40-year-old male who complained of fever for 1 day before being confirmed as having COVID-19 by PCR. Although his case never progressed to a more severe condition, low grade fever continued for a prolonged period. In agreement with this patient’s clinical course, PCRs were repeatedly performed from 19 to 34 days post-onset, and they showed positive and negative results alternately.

**Fig. 1.**
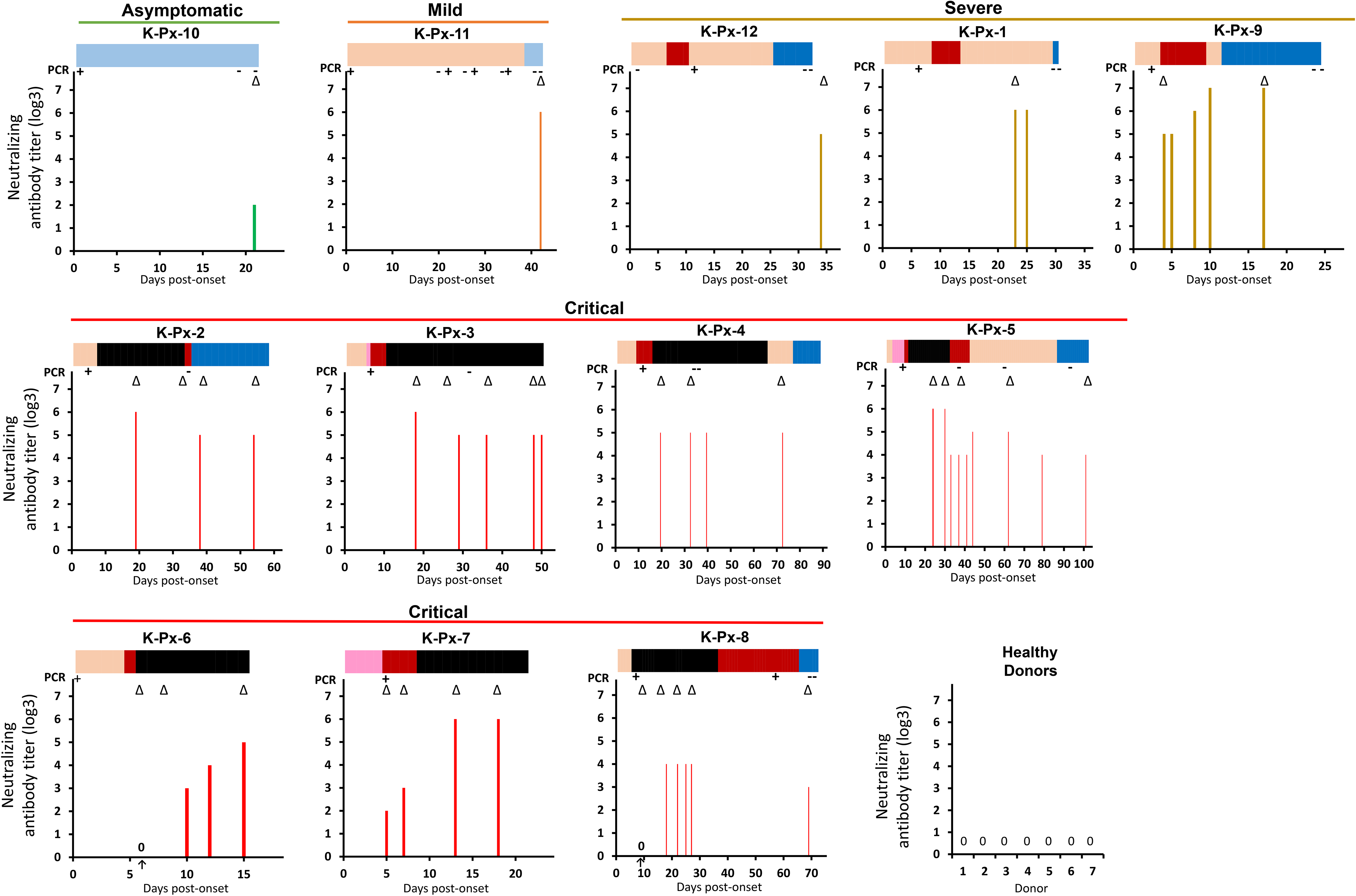
Neutralizing antibody titer in the COVID-19 patients’ sera. Neutralizing antibody titers against SARS-CoV-2 were analyzed. Each panel is for the indicated patient and includes results for the sera obtained at single or several time point(s) from admission to hospital until discharge. The bars on top of each panel indicate the severity of symptoms in chronological order. *Light Blue:* asymptomatic. *Orange:* upper respiratory symptoms without pneumonia. *Pink:* pneumonia requiring no supplemental oxygen. *Red:* requiring supplemental oxygen. *Black:* requiring intensive care including mechanical ventilation. *Dark Blue:* Getting better with residual symptoms. The results of the neutralization assay of the control sera from seven healthy donors are also shown. Number 0 in the panel indicates no detection of neutralizing activity. Δ below the bar indicates time point(s) for cytokines/chemokines measurement showed at Fig. 2.

Our analysis of the induction of neutralizing antibody during the first days after disease onset in patients K-Px-6, -7, -8 and -9 revealed that neutralizing antibody started to be induced around day 4-10 post-onset, and titer then gradually increased, indicating that seroconversion had occurred around these days. To elucidate the effect of neutralizing antibody production on the persistence of virus replication, we investigated the time point(s) at which neutralizing antibody started to be detected and virus genome became undetected. In all of the severe/critical patients (excluding the two who died in which a follow-up PCR was not performed), neutralizing antibody had already been induced at the time that virus genome became undetected by PCR. A different pattern was observed in patient K-Px-8, in whom viral genome could still be detected at day 56 post-onset even though neutralizing antibody had already been induced. Of note, K-Px-8 is the only critical patient in this study who received corticosteroid pulse therapy from the beginning of the disease course.

We thus observed that all the individuals infected with SARS-CoV-2 developed neutralizing antibody, including the asymptomatic patient (although the titer was low).

### The trend of cytokine/chemokine expression in patients with different COVID-19 severities

Cytokine release has been shown to be related to COVID-19 severity [8, 9]. We therefore measured the levels of 48 different cytokines, chemokines, and growth factors (Table 1) in sera of COVID-19 patients with different severities, at various time points. Figure 2 provides the levels of the measured cytokines, chemokines, and growth factors in patients’ sera, expressed in picograms per milliliter of serum. In order to more easily analyze the trend of the individual cytokine expressions in the different disease-severity groups, we have presented the measured value of each cytokine or chemokine in a graded color scale (heat map). The average measured values for each cytokine or chemokine in two healthy controls were also calculated, and the values higher than those control values are scaled red-colored according to the amount.

**Fig. 2.**
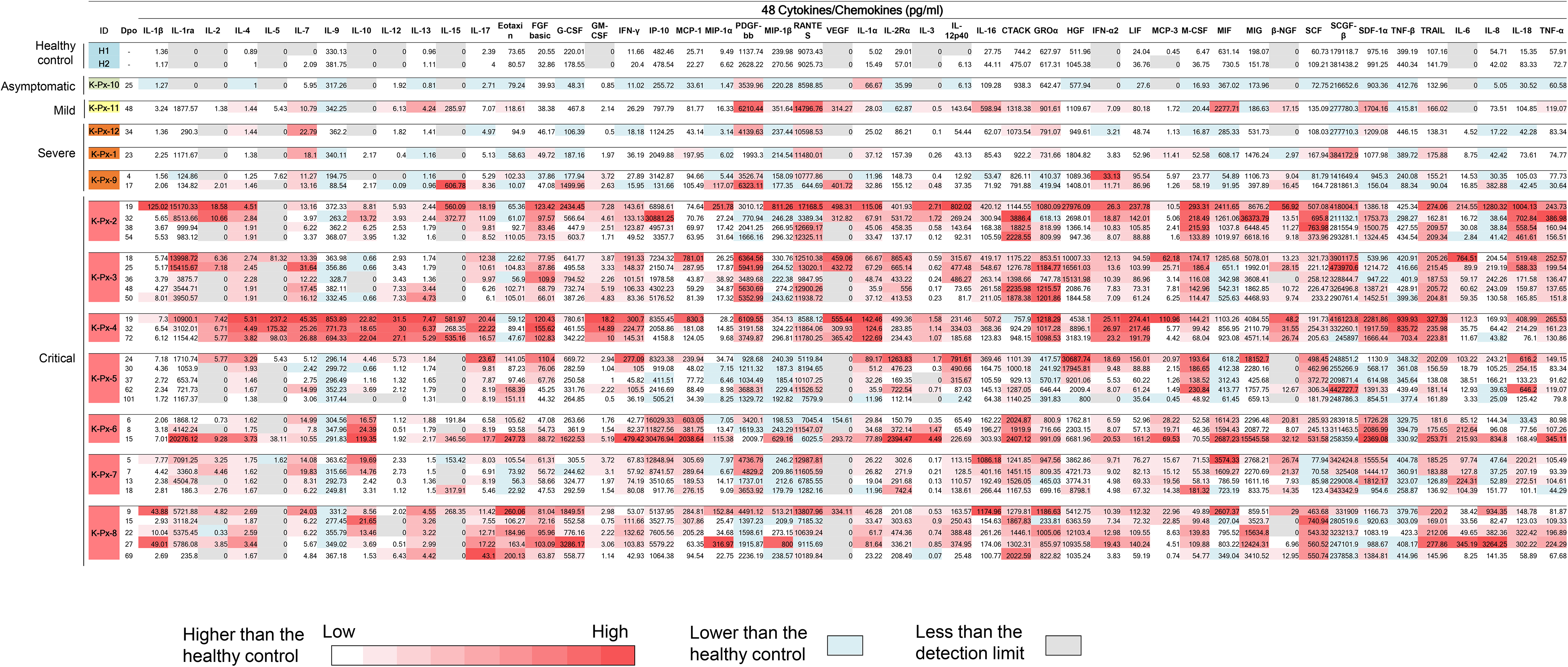
Cytokine and chemokine levels in the COVID-19 patients’ sera. The concentrations of 48 cytokines and chemokines in sera were measured by the multiplex suspension array system with the Bio-plex Pro™ human cytokine screening 48-plex panel (BioRad). The blank-corrected observed values were translated to the concentration with the standard curve for each cytokine or chemokine and further corrected by the dilution rate. The average values of duplicate wells are shown. Values less than the detection limit were treated as Zero and colored *gray*. The average value of the two healthy controls was calculated for each cytokine or chemokine, and values lower than the average are *pale blue*. Values higher than the average are *scaled-red* according to the amount.

Six of the seven critical patients required mechanical ventilation support after several days of hospitalization, suggesting that disease progressivity continued during treatment. The day of ventilator introduction may thus reflect the time point at which the disease clinically progresses to a more severe condition, and the day of ventilator removal is likely to reflect the time point at which patient shows sufficient clinical improvement in respiratory function. We thus analyzed the kinetics of cytokine/chemokine release in relation to the patients’ clinical presentations. As shown in patient K-Px-2, the levels of several cytokines and chemokines were higher during his ventilator usage period (days 8–33 post-onset) — especially in the earlier days □ — □ compared to levels beyond this period. A similar trend was observed in other critical COVID-19 patients who recovered (K-Px4, K-Px-5, and K-Px-8), with the exception of patient K-Px-4, whose cytokine and chemokine levels were constantly high for an extended period of time including after recovery period.

For three critical COVID-19 patients who died (K-Px-3, K-Px-6, and K-Px-7), the trend of cytokine/chemokine release was unique to each patient. Patient K-Px-3 showed consistently high levels of most cytokines and chemokines from 1 week after the start of ventilator use until death (day 18 until day 50 post-onset) with the following exceptions, which showed a reducing trend toward death: IL-1ra, IL-5, IL-17, MCP-1, VEGF, IL-12p40, IL-16, HGF, IFN-α2, MCP-3, IL-6, IL-18, and TNF-α.

A different pattern was observed in patient K-Px-6, with a clearly increased level of most cytokines and chemokines in the days before death, emphasizing that this ‘cytokine storm’ was the possible cause of death. Interestingly, patient K-Px-7, who had more comorbidities related to COVID-19 severity compared to the other patients, did not show an increasing trend of cytokines or chemokines toward death.

## DISCUSSION

We analyzed the trend of neutralizing antibody production against SARS-CoV-2 and cytokine/chemokine levels in a small series of COVID-19 patients with different severities. We observed a clear neutralizing antibody response against SARS-CoV-2 in all 12 patients, whereas healthy controls did not show any neutralizing response at all, suggesting a highly specific nature of neutralizing antibody against SARS-CoV-2. Critically and severely infected patients showed higher neutralizing activity for SARS-CoV-2 compared to asymptomatic patient. This result is in agreement with two reports that SARS-CoV-2-specific neutralizing antibody titer was higher in patients with severe infections compared to those with mild infections [6, 7]. In the present asymptomatic patient, a few increases in neutralizing antibody titer were seen, which may indicate that virus replication was not very high or that it can replicate but only at local sites without producing any symptoms. These results indicate that virus replication in severe cases occurs in the lungs or systemically, and that in asymptomatic cases the replication is not very high.

Despite being categorized as having a critical COVID-19 infection, patient K-Px-8 showed a relatively low neutralizing antibody titer against SARS-CoV-2 on multiple measurements at different time points. This may be related to the use of methylprednisolone pulse therapy from a relatively early phase of the disease (day 6 post-onset), at which time neutralizing antibody may not yet be induced.

Although patient K-Px-11 was considered to have a mild infection, he continuously presented with prolonged low grade fever, and serial viral genome detections showed positive and negative results alternately, suggesting a continued replication of the virus for a relatively long period. Therefore, the relatively high titer of neutralizing antibody detected in this patient is not a surprising result.

By analyzing the trend of neutralizing antibody titer in these severe and critical COVID-19 patients, we observed that neutralizing antibody against SARS-CoV-2 seems to be induced around 4-10 days after disease onset. This observation is similar to that of a previous report of a young man with a mild SARS-CoV-2 infection in whom neutralizing antibody titer rapidly increased from day 4 until day 20 [18]. Additionally, another report showed neutralizing antibody against SARS-CoV-2 could be detected from day 10-15 after onset [19]..

Not surprisingly, we also observed that neutralizing antibody was always induced before SARS-CoV-2 genome testing showed a negative result, suggesting efficient viral clearance by a successful induction of neutralizing antibodies. In addition, decreasing titer of neutralizing antibody detected on day 33 in patient K-Px-5 was accompanied by clinical improvement, as evident by ventilator weaning on day 32.

Regarding the assay of cytokine/chemokine release, our data showed clearly higher amounts of most cytokines and chemokines in critical COVID-19 patients compared to patients with milder symptoms (Fig. 2) at all time points. This observation is in agreement with reports that cytokine storm is associated with COVID-19 severity [20].

IL-6, which plays a central role in the cytokine storm [9], signals through two main pathways (cis- and trans-signaling pathways) which contribute to CRS [8, 21]. Consistent with earlier reports, our present analyses demonstrated that serum IL-6 level was clearly increased in all 10 of the severe/critical COVID-19 patients, but not in asymptomatic and mild patients. However, our results indicated that determinant of disease severity was not only the induction of IL-6, but also the induction of the other cytokines and/or chemokines.

Although most of the cytokines and chemokines tested herein were high in critical COVID-19 patients compared to those with milder symptoms, some cytokines and/or chemokines were detected at higher levels in some of the patients with milder symptoms. Among these cytokines/chemokines were PDGF-bb, RANTES, IFN-α2, MIF, SCGF-β, and SDF-1α. This result is inconsistent with a report that PDGF-BB, RANTES, and IL-9 were consistently low in the group of patients with severe and fatal infections compared to the mild-infection group [22]. Our data showed that RANTES and PDGF-BB were not always higher in the non-severe patients compared to the severe patients, whereas IL-9 was always detected at values that were comparable to those of the healthy controls in all patients regardless of disease severity, except for patient K-Px-4 who consistently showed relatively high levels of most of the cytokines and chemokines over an extended period of time, including the recovery phase.

Based on the results of our analyses of the courses of COVID-19 patients’ clinical conditions, neutralizing antibody titers and cytokine/chemokine levels, we suggest the following concept. In severe COVID-19 patients, SARS-CoV-2 replicates abundantly, especially in the lung and/or systemically. The higher positivity rate of SARS-CoV-2 detection in sputum compared to nasopharyngeal or throat swabs may be explained by the more efficient replication of this virus in the lung [23–26]. Innate immunity is then induced in response to viral replication, which is then followed by the induction of acquired immunity as shown by the detection of specific antibody and T lymphocytes recognizing SARS-CoV-2. The induction of innate immunity is reflected by the increased levels of cytokines and chemokines in serum, which are responsible for the cytokine storm. Both virus replication and cytokine storm contribute to the lung damage that occurs in patients with severe disease.

The induction of acquired immunity may then efficiently clear the virus, which is evidenced by our observation that at the time the neutralizing antibody reached its higher level, viral genomes turned undetected. However, by the time acquired immunity is induced, the disease may still have been progressing continuously and lung damage continues as the cytokine storm has not been controlled.

Based on this understanding, we suggest that convalescent plasma therapy from COVID-19 recovered patients to severely infected patients, human or humanized antibody therapy, or effective antiviral agent (when available) may provide better outcomes when administered earlier in the disease process before severe cytokine production begins. Once viral infection is eliminated in severe cases, steroid therapy may be added to suppress the cytokine storm.

We used authentic SARS-CoV-2 to measure neutralizing activity in this study. Recent studies generally used a pseudotyped virus, for the neutralizing assay[11, 12]. However there were inconsistent results in neutralizing assay between pseudotyped and authentic viruses [13–15], which may be caused by different presentation of S protein resulting from different environmental condition [13]. Therefore, it seems that the authentic virus should be used to examine the neutralizing activity against SARS-CoV-2.

The neutralizing antibody titer for SARS-CoV-2 was recently shown to decrease gradually [27]. Our present findings demonstrated that in patients with more severe infections, neutralizing titer is relatively high, and therefore patients’ antibodies may be retained for a long period. It will be informative to continue measuring these patients’ neutralizing antibody titers and to examine whether the antibodies can protect patients from a second infection of SARS-CoV-2.

## Data Availability

NO data is available.

## Acknowledgement

We thank Kazuro Sugimura MD, PhD (Executive Vice President, Kobe University) for his full support to promote this study. We also thank Yukiya Kurahashi, Zhenxiao Ren, Anna Lystia Poetranto, Salma Aktar, Jing Rin Huang, and Silvia Sutandhio (Division of Clinical Virology, Center for Infectious Diseases, Kobe University Graduate School of Medicine) for technical support in this study. We express our sincere gratitude for cooperation and participation of staffs of Hyogo Prefectural Kakogawa Medical Center. We thank Research Foundation for Microbial Diseases of Osaka University (BIKEN), Osaka University for providing SARS-CoV-2 strain.

## Author Contributions

L.T and T.N equally contributed to this work.

Concept and design: L.T, T.N, Y.M.

Acquisition, analysis, or interpretation of data: L.T, T.N, S.F, S.I, K.F, M.N, J.A, Y.N, Y.M

Drafting of the manuscript: L.T, Y.M

Critical revision of the manuscript for important intellectual content: L.T, T.N, Y.M

Administrative, technical, or material support: S.F, K.F, M.N, J.A

Supervision: Y.N, Y.M

## Summary

Neutralizing antibody against SARS-CoV-2 is induced at high titer in severe COVID-19 patients, followed by termination of virus replication while continuously high cytokines/chemokines. Convalescent plasma therapy could be administered early to eliminate virus, followed by corticosteroid after viral genome disappearance.

